# Participatory research in health intervention studies involving migrants: a systematic review

**DOI:** 10.1101/2021.06.09.21258458

**Authors:** Kieran Rustage, Alison Crawshaw, Saliha Majeed-Hajaj, Anna Deal, Laura B Nellums, Yusuf Ciftci, Sebastian Fuller, Lucy Goldsmith, Jon S Friedland, Sally Hargreaves

## Abstract

**Objective:** To analyse the use of participatory approaches in research of health interventions for migrants, and how utilised approaches embody core participatory principles of democracy and power-distribution.

**Design:** A systematic review of original articles. Electronic searchers were carried out in the databases MEDLINE, Embase, Global Health and PsychINFO (from inception – Nov 2020).

**Eligibility criteria for study selection:** The analysis included original peer-reviewed research which reported on attempts to develop and implement a health intervention for migrants using participatory approaches. We defined migrants as foreign-born individuals; studies using definitions demonstrably outside of this were excluded. Only articles reporting the full research cycle (inception, design, implementation, analysis, evaluation, dissemination) were included.

**Data extraction:** Information related to who was involved in research (migrants or other non-academic stakeholders), the research stage at which they were involved (inception, design, implementation, analysis, evaluation, dissemination), and how this involvement aligned with the core principles of participatory research – categorising studies as exhibiting active, pseudo, or indirect participation of migrants.

**Results:** 1793 publication were screened of which 28 were included in our analysis. We found substantial variation in the application of participatory research approaches: across 168 individual research stages analysed across the 28 studies, we recorded 46 instances of active participation of migrants; 30 instances of proxy participation; and 24 instances of indirect participation. Whilst all studies involved at least one non-academic stakeholder group in at least one stage of the research, just two studies exhibited evidence of active participation of migrants across all research stages.

**Conclusions:** These data highlight important shortfalls in the inclusion of migrant groups in developing health interventions that affect their lives and suggest a more rigorous and standardised approach to defining and delivering participatory research is urgently needed to improve the quality of participatory research.

**Registration:** This review followed PRISMA guidelines and is registered on the Open Science Framework (osf.io/2bnz5)

**Strengths and Limitations:**

- This systematic review represents a robust and novel assessment of the applications of participatory approaches and principles to health intervention research with migrants.
- This review casts a critical lens over the application and outcomes of participatory approaches, conceptually focusing on the relationship between the methods used and the populations involved, and how this all relates to participatory principles.
- This review is limited by the varied and interchangeable use of participatory research terms within this field. The categorisations and terms we introduce may therefore be defined differently by others.
- This review is limited by the lack of clear and consistent reporting of participatory methods utilised; guidelines must be prepared and consistently adopted to improve transparency in all participatory research.
- This does not analyse or consider associations between participatory methods, and health or research outcomes; future research to understand any such associations is needed.

## Introduction

Considerable emphasis is now being placed on ensuring patient and public engagement in health research, including striving for greater involvement of marginalised groups such as migrants and ethnic minorities.^1,2^ However, whether this is effectively and meaningfully done in practice to ensure truly patient-centred research has yet to be fully elucidated. Participatory research represents a distinct research paradigm in which research is done collaboratively the individuals whose lived experiences and actions are the subject of study, as active partners who share power and influence over research processes and outcomes. ^3-6^

The fundamental principles of participatory research are those of inclusivity and democracy, particularly in relation to those directly affected by the research in question. ^3^ Included under the umbrella of participatory research approaches are more specific methodologies which look to uphold these principles, including: community based participatory research (CBPR); action research; some patient & public involvement (PPI); as well as broader derivatives such as community-based collaborative action research. Participatory research holds the potential to bridge the gap between public health research and practice, creating a context in which patients and the public have meaningful influence over research decisions, increasing the relevance and impact of research outcomes to their own lives. ^7^

Participatory research is likely to be particularly powerful when working with underserved and marginalised groups such as migrants, where traditional research has frequently failed to provide an appreciable health benefit. Whilst a heterogenous group, comprising a multitude of cultures, ethnicities and socio-cultural circumstances, many migrants can find themselves in vulnerable situations, marginalised by health systems, ^8,9^ and society alike. ^10,11^ There is a growing consensus around the need for academics and health systems to become more responsive to, and inclusive of refugee and migrant health concerns, ^12^ with the ultimate goal to collaboratively pursue research that is better centred around, and grounded in the needs of migrant communities. Indeed, community engagement in public health interventions has already been shown to be effective when working with marginalised groups for a range of health outcomes, and can provide benefits to participants themselves, such as in improving health behaviours and participant self-efficacy. ^13^

Despite the potential of participatory research, there are varied interpretations as to how to apply such approaches. Challenges exist in deciding who should be involved, and whether, involvement should extend beyond the target group (for example migrants), to other non-academic stakeholder groups such as community organisations and professionals. ^3^ There are also differing interpretations of the degree of participation required of individuals for research to be considered participatory rather than tokenistic, though it is suggested that unless involved individuals are partners or co-researchers throughout the entirety of a project, the work cannot be participatory. ^3,14^ Overall, it is widely agreed quality participation is characterised by non-academic stakeholders having opportunities to engage in research, make decisions, and perform leadership roles around such research, ^5^ empowering the public at the highest level and asserting their right to be involved in decision-making and to influence outcomes. Understanding the different approaches to participatory research and whether the core principles of participatory research are upheld is crucially important if good practice is to be identified.

We therefore did a systematic review to analyse participatory approaches used in health intervention research focused on migrants. We explored at what stages, and how migrants (or other non-academic stakeholders) were involved and analysed how this compares to principles of participatory research. We also looked for evidence of how such approaches had been evaluated by study authors, and how effective it was assessed to be.

## Methods

We did a systematic review, following PRISMA guidelines, which is registered on the Open Science Framework (osf.io/2bnz5). The primary aim of this systematic review was to describe and analyse the participatory approaches utilised in health intervention research in which the intervention was aimed at benefitting migrant populations. We specifically sought to identify evidence on which stages of the research (inception, design, implementation, analysis, evaluation, dissemination) migrants (or other non-academic stakeholders) were involved in. We then analysed how they were involved and what influence they had over the research processes, comparing this to core principles of participatory research. Secondary outcomes were related to describing author evaluations of the impact utilising a participatory approach had on the research process, or of the participatory approach itself.

### Inclusion and Exclusion criteria

We included peer-reviewed primary-research reporting on health interventions aimed at benefitting migrant populations that described using a participatory approach across the whole research process. Research which purported to use a participatory research approach through descriptors in their introduction and methods, or which utilised a recognised participatory approach such as community-based participatory research (CBPR), action research, or community-based collaborative action research, and specifically targeted migrants, was included in the review. We defined migrants as foreign-born individuals and considered a health intervention to be any initiative, tool, or programme that looked to improve health outcomes, including those related to mental health and health literacy.

Studies were excluded if they did not report on all stages of research into the health intervention: Inception, design, implementation, analysis, evaluation, dissemination.

As such, publications presenting interim results of studies which had not completed the full research cycle, as well as studies specifically focusing on only co-designing interventions were excluded. We took this approach so as not to unfairly penalise ongoing research in our analysis, nor co-designed research; we consider co-design to be one component of the broader participatory research paradigm and were most interested in how approaches manifest across the entirety of a research cycle.

Studies explicitly defining migrant status according to ethnic or ancestral background but not country of birth were excluded, as were papers where primary data were not reported (e.g., comments, editorials, letters, and reviews).

### Search strategy

We searched the databases MEDLINE, Embase, Global Health and PsychINFO from inception – November 2019 utilising a Boolean search strategy with keywords and medical subheadings (MeSH) related to two major themes: migrants and participatory research. There were no geographic or language restrictions. An additional text file shows a representative search strategy (see Additional file 1). We identified additional studies through hand searching the bibliographies of publications included after full-text screening.

### Study selection and quality assessment

Two reviewers duplicated the title and abstract screening and full-text screening (KR & SM-H), which was carried out using the web-based application Rayyan. ^15^ The reasons for excluding studies during full-text screening were recorded. Any discrepancies in screening decision between the two initial reviewers were mediated by a third reviewer (AC).

### Data extraction and analysis

Studies which reported using participatory research approaches, and which reported on all stages of the research were extracted using a piloted form by KR and SH.

We extracted summary data on geographical location, the self-described participatory approach, specific target population, and aims of the research. Further data relating to the participatory approach of each study was extracted, categorised, and analysed.

We extracted data on the stages of the research in which migrants were involved. Where specific research stages did not involve migrants, but did involve other non-academic stakeholders this was recorded, sub-categorising these groups as community groups/third-sector organisations or professional services. We subsequently extracted data on the methods used to involve migrants (or other non-academic stakeholders) at each stage of the research (inception, design, implementation, analysis, evaluation, and dissemination). An additional file provides details of the summary extracted data (see Additional file 2).

Using this information, we analysed the overall participatory character of the studies from the perspective of inclusion and decision-making power of migrants specifically. For each stage of the research, we categorised these data in one of three ways (active, pseudo, indirect) within a framework developed with reference to the literature, particularly that relating to participatory research as a democratic process and being necessary to implement at all stage of the research. ^3,5,14,16^ The final framework and definitions were agreed by all co-authors (Table 1).

**Table 1.** Classifications and definitions of participatory research utilised in this systematic review.

| Category of Participation |  | Definition |
| --- | --- | --- |
| Active Participation |  | Evidence migrants (as the target population) were actively involved and wielded influence in decisions relating to the research (e.g., involved in voting on decisions, or being participants in steering committees which made the decisions). |
| Pseudo-participation | Proxy-participation | Evidence community groups/third-sector organisations and/or professional services were actively involved and wielded influence in decisions relating the research. However, migrants (as the target population) did not appear to be actively involved or were indirect participants at most. |
|  | Indirect participation | Evidence migrants (as the target population) were subject to research activities (e.g., surveys, interviews), or partook in implementing/actioning research directions (e.g., delivering the designed intervention), but did not appear to have any active involvement or influence in decisions relating to this stage of the research. Furthermore, there was no evidence of community group/third-sector organisation involvement or influence. |
| No explicit evidence |  | There was no explicit evidence of the participation of migrants, community groups/third-sector organisations, or professional services regarding decision making processes related to the research stage. |

### Patient and Public Involvement

Members of our authorship team have past and current experience of working within third sector organisations. This experience helped to frame the research questions, and definitions used in the analysis. However, lay-patients and public specifically were not involved in this research.

## Results

### Screening results

Database searches returned 1793 results; a total of 292 duplicates were removed, and 1501 publications were retained for title and abstract screening. Of the 1501 remaining publications, 1357 were excluded during title and abstract screening, and 144 were retained for full-text screening. During full-text screening, 116 publications did not meet our criteria and were excluded, with the reasons for exclusion recorded (Fig 1.). Overall, 28 publications met the inclusion criteria and were included in this systematic review (Table 2.).

**Table 2.**
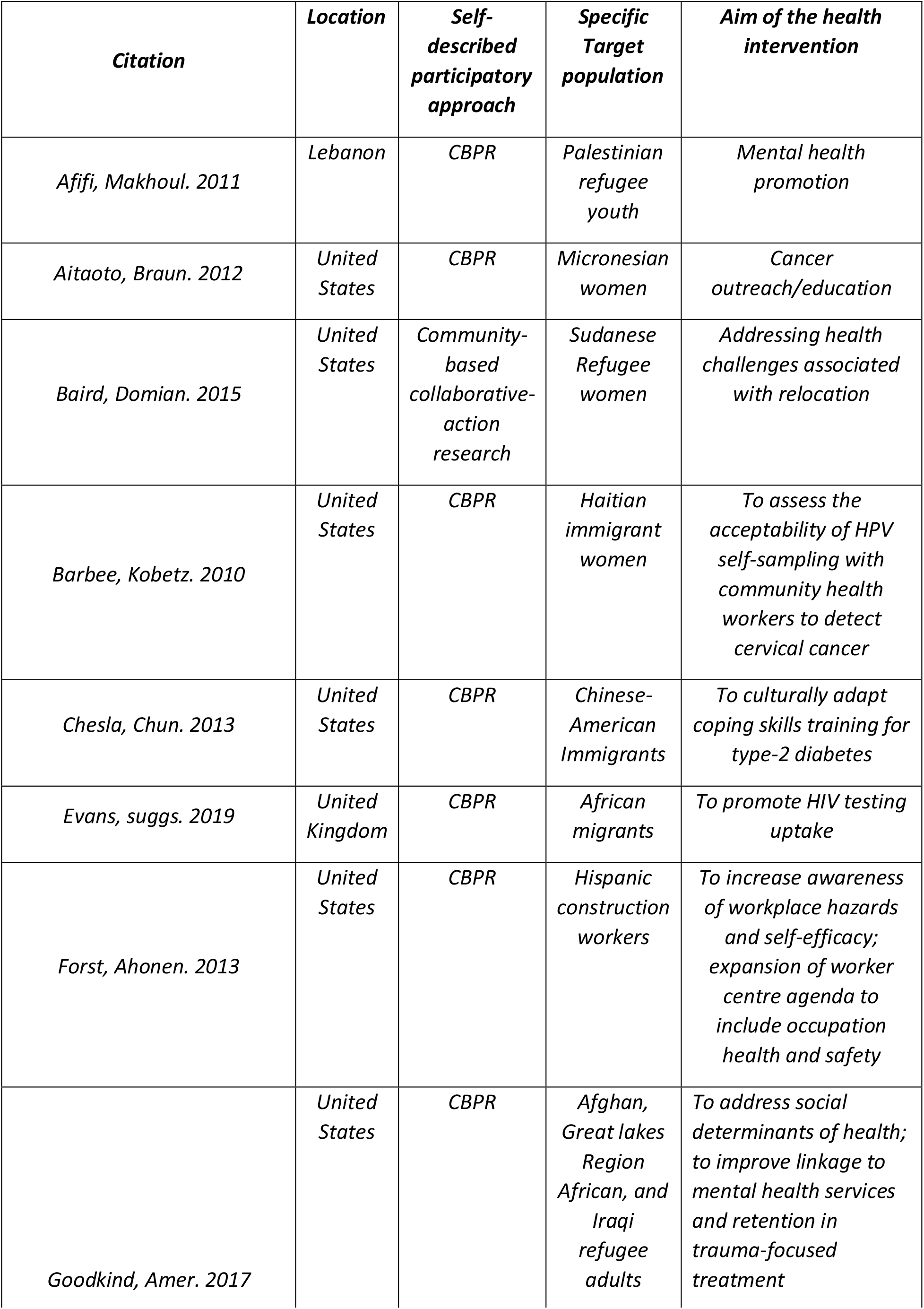
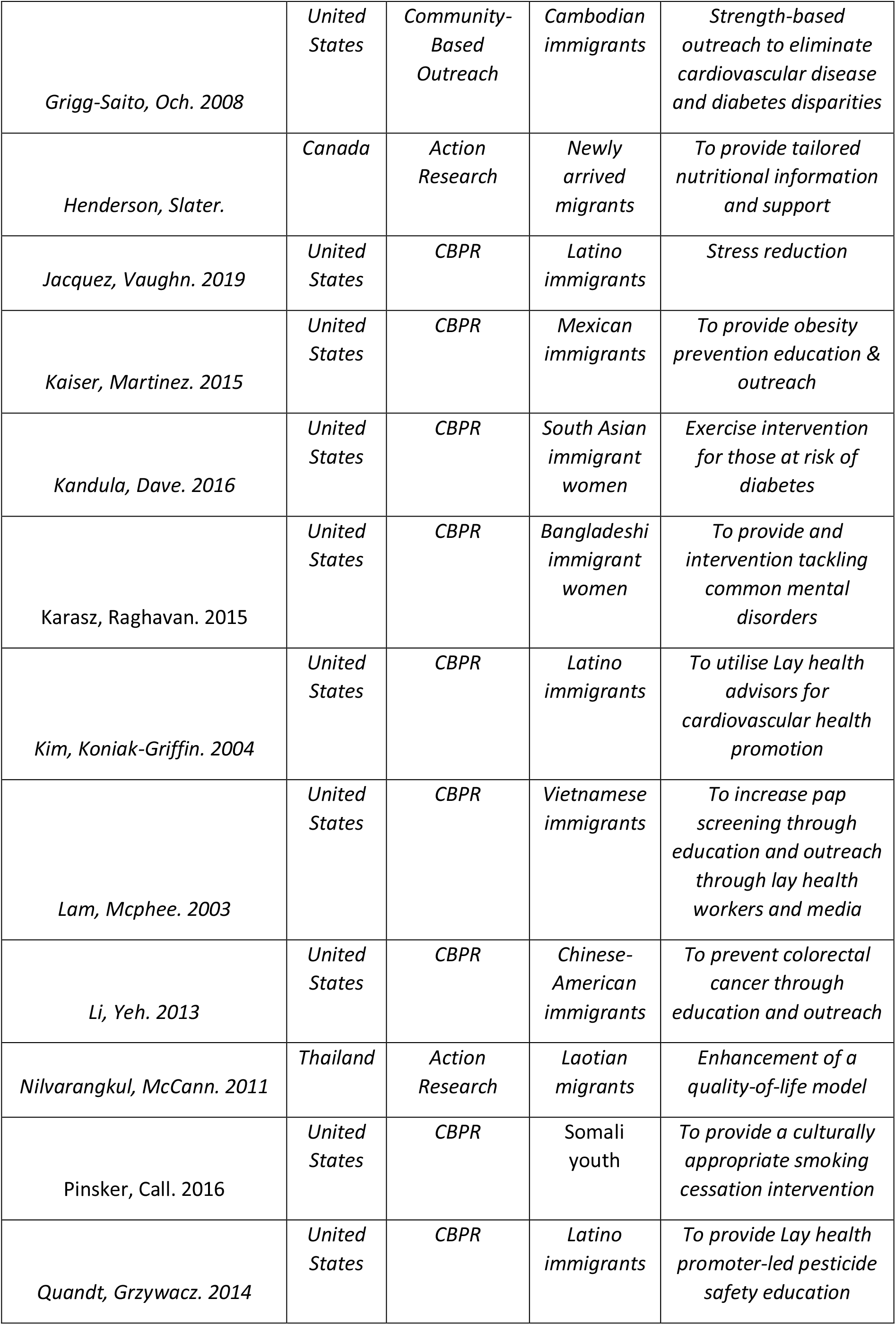
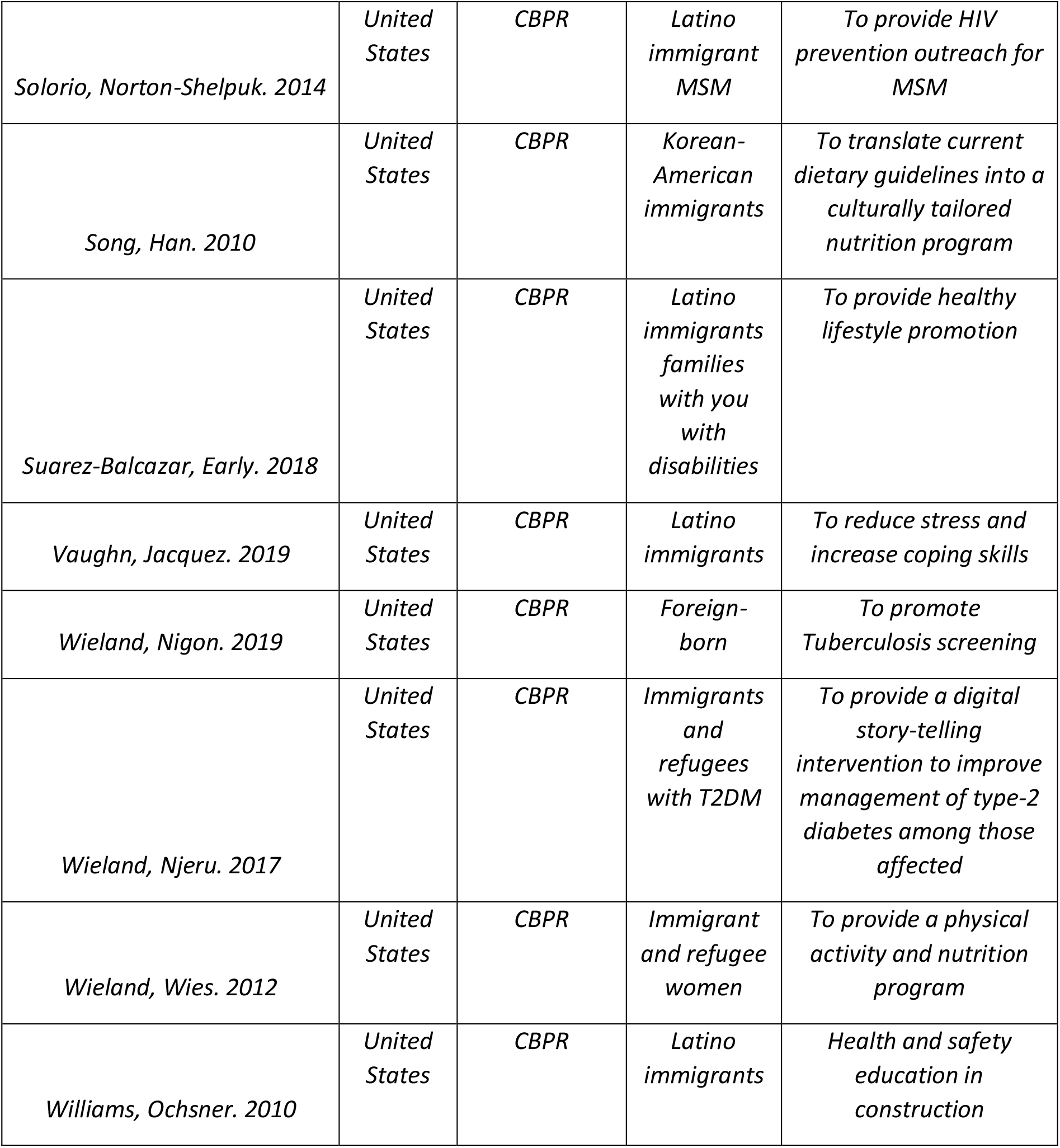
Descriptive characteristic of studies included in this systematic review.

| ***Citation*** | ***Location*** | ***Self-described participatory approach*** | ***Specific Target population*** | ***Aim of the health intervention*** |
| --- | --- | --- | --- | --- |
| *Afifi, Makhoul. 2011* | *Lebanon* | *CBPR* | *Palestinian refugee youth* | *Mental health promotion* |
| *Aitaoto, Braun. 2012* | *United States* | *CBPR* | *Micronesian women* | *Cancer outreach/education* |
| *Baird, Domian. 2015* | *United States* | *Community-based collaborative-action research* | *Sudanese Refugee women* | *Addressing health challenges associated with relocation* |
| *Barbee, Kobetz. 2010* | *United States* | *CBPR* | *Haitian immigrant women* | *To assess the acceptability of HPV self-sampling with community health workers to detect cervical cancer* |
| *Chesla, Chun. 2013* | *United States* | *CBPR* | *Chinese-American Immigrants* | *To culturally adapt coping skills training for type-2 diabetes* |
| *Evans, suggs. 2019* | *United Kingdom* | *CBPR* | *African migrants* | *To promote HIV testing uptake* |
| *Forst, Ahonen. 2013* | *United States* | *CBPR* | *Hispanic construction workers* | *To increase awareness of workplace hazards and self-efficacy; expansion of worker centre agenda to include occupation health and safety* |
| *Goodkind, Amer. 2017* | *United States* | *CBPR* | *Afghan, Great lakes Region African, and Iraqi refugee adults* | *To address social determinants of health; to improve linkage to mental health services and retention in trauma-focused treatment* |
| *Grigg-Saito, Och. 2008* | *United States* | *Community-Based Outreach* | *Cambodian immigrants* | *Strength-based outreach to eliminate cardiovascular disease and diabetes disparities* |
| *Henderson, Slater.* | *Canada* | *Action Research* | *Newly arrived migrants* | *To provide tailored nutritional information and support* |
| *Jacquez, Vaughn. 2019* | *United States* | *CBPR* | *Latino immigrants* | *Stress reduction* |
| *Kaiser, Martinez. 2015* | *United States* | *CBPR* | *Mexican immigrants* | *To provide obesity prevention education & outreach* |
| *Kandula, Dave. 2016* | *United States* | *CBPR* | *South Asian immigrant women* | *Exercise intervention for those at risk of diabetes* |
| *Karasz, Raghavan. 2015* | *United States* | *CBPR* | *Bangladeshi immigrant women* | *To provide and intervention tackling common mental disorders* |
| *Kim, Koniak-Griffin. 2004* | *United States* | *CBPR* | *Latino immigrants* | *To utilise Lay health advisors for cardiovascular health promotion* |
| *Lam, Mcphee. 2003* | *United States* | *CBPR* | *Vietnamese immigrants* | *To increase pap screening through education and outreach through lay health workers and media* |
| *Li, Yeh. 2013* | *United States* | *CBPR* | *Chinese-American immigrants* | *To prevent colorectal cancer through education and outreach* |
| *Nilvarangkul, McCann. 2011* | *Thailand* | *Action Research* | *Laotian migrants* | *Enhancement of a quality-of-life model* |
| *Pinsker, Call. 2016* | *United States* | *CBPR* | *Somali youth* | *To provide a culturally appropriate smoking cessation intervention* |
| *Quandt, Grzywacz. 2014* | *United States* | *CBPR* | *Latino immigrants* | *To provide Lay health promoter-led pesticide safety education* |
| *Solorio, Norton-Shelpuk. 2014* | *United States* | *CBPR* | *Latino immigrant MSM* | *To provide HIV prevention outreach for MSM* |
| *Song, Han. 2010* | *United States* | *CBPR* | *Korean-American immigrants* | *To translate current dietary guidelines into a culturally tailored nutrition program* |
| *Suarez-Balcazar, Early. 2018* | *United States* | *CBPR* | *Latino immigrants families with you with disabilities* | *To provide healthy lifestyle promotion* |
| *Vaughn, Jacquez. 2019* | *United States* | *CBPR* | *Latino immigrants* | *To reduce stress and increase coping skills* |
| *Wieland, Nigon. 2019* | *United States* | *CBPR* | *Foreign-born* | *To promote Tuberculosis screening* |
| *Wieland, Njeru. 2017* | *United States* | *CBPR* | *Immigrants and refugees with T2DM* | *To provide a digital story-telling intervention to improve management of type-2 diabetes among those affected* |
| *Wieland, Wies. 2012* | *United States* | *CBPR* | *Immigrant and refugee women* | *To provide a physical activity and nutrition program* |
| *Williams, Ochsner. 2010* | *United States* | *CBPR* | *Latino immigrants* | *Health and safety education in construction* |

**Figure 1.**
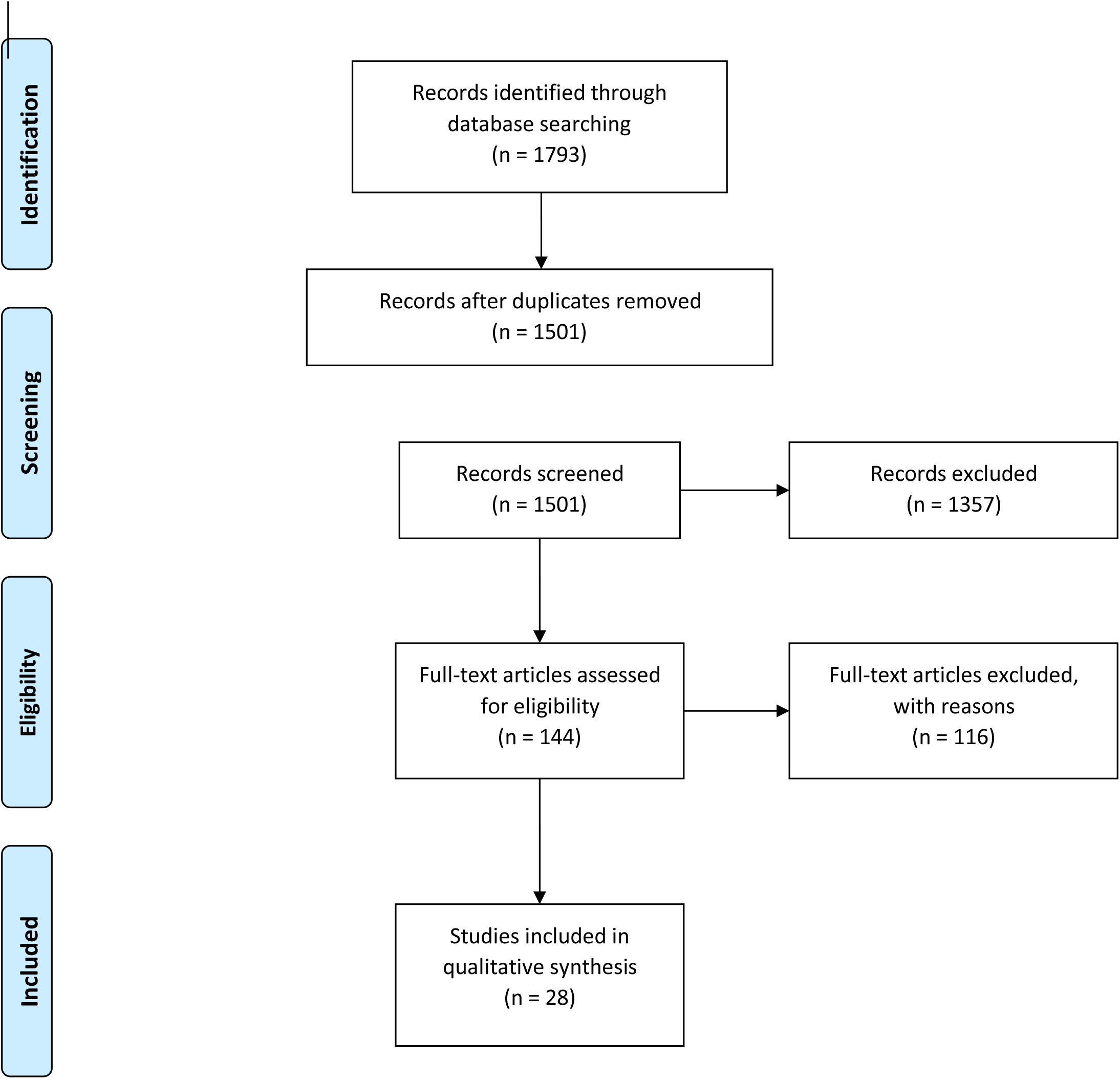
PRISMA flow-diagram of the study selection process.

### Study Characteristics

The research articles included in this systematic review were published between 2003 and 2019. Only 13 of the publications had any discernible dates relating to when the reported work was conducted, with these dates being between 2003 and 2018. The majority of the publications related to work carried out in the United States (24 out of 28); the remaining publications related to work carried out in Canada, ^17^ Lebanon, ^18^ Thailand, ^19^ and the United Kingdom. ^20^ The self-described approach taken by 24 of the 28 included studies was community-based participatory research (CBPR) ^18,20-42^; the remaining four studies described using community-based collaborative-action research, ^43^ community-based outreach, ^44^ and action research. ^17,19^. The dominant focus of the included studies was around education or outreach (Table 2), for example around cancer education, ^21^ or healthy lifestyles promotion; ^37^ five studies specifically mentioned including refugees (Table 2).^18,25,40,41,43^

### Analysis of participation within included studies

In our analysis, participation varied substantially according to the stage of the research under scrutiny. Only two of the included studies reported explicit evidence of some degree of participation of at least one non-academic stakeholder groups across all research stages.^25,26^ Overall, we extracted and categorised evidence of the participation of at least one non-academic stakeholder group in 22 studies during the inception; ^17-19,21,22,25-30,32,34-36,38-44^ 25 studies during the design; ^17-19,21-35,37-42,44^ 23 studies during implementation; ^19-28,30,31,33-39,41-44^ 4 studies during analysis; ^25,26,35,39^ 22 studies during evaluation; ^17,19,20,23-33,35-38,40,42-44^ and 4 studies during dissemination (Table 3.). ^22,25,26,38^

**Table 3.** Analysis and categorisation of participatory character displayed across research stages within included studies

| *Citation* | *Research Stage* |  |  |  |  |  |
| --- | --- | --- | --- | --- | --- | --- |
|  | *Inception* | *Design* | *Implementation* | *Analysis* | *Evaluation* | *Dissemination* |
| *Afifi, Makhoul. 2011* | ○ | ◆ | X | X | X | X |
| *Aitaoto, Braun. 2012* | ● | ◆ | ◆ | X | X | X |
| *Baird, Domian. 2015* | ◆ | X | ○ | X | ○ | X |
| *Barbee, Kobetz. 2010* | ◆ | ◆ | ◆ | X | X | ◆ |
| *Chesla, Chun. 2013* | X | ◆ | ● | X | ○ | X |
| *Evans, suggs. 2019* | X | X | ○ | X | ○ | X |
| *Forst, Ahonen. 2013* | X | ● | ● | X | ◆ | X |
| *Goodkind, Amer. 2017* | ◆ | ◆ | ◆ | ◆ | ◆ | ◆ |
| *Grigg-Saito, Och. 2008* | ◆ | ◆ | ◆ | X | ◆ | X |
| *Henderson, Slater.* | ● | ○ | X | X | ○ | X |
| *Jacquez, Vaughn. 2019* | ◆ | ◆ | ◆ | ◆ | ◆ | ◆ |
| *Kaiser, Martinez. 2015* | ○ | ◆ | ○ | X | ○ | X |
| *Kandula, Dave. 2016* | ● | ● | ● | X | ● | X |
| *Karasz, Raghavan. 2015* | ● | ◆ | X | X | ○ | X |
| *Kim, Koniak-Griffin. 2004* | ● | ◆ | ◆ | X | ◆ | X |
| *Lam, Mcphee. 2003* | X | ◆ | ◆ | X | ◆ | X |
| *Li, Yeh. 2013* | ● | ● | X | X | ○ | X |
| *Nilvarangkul, McCann. 2011* | ◆ | ◆ | ● | X | ● | X |
| *Pinsker, Call. 2016* | X | ◆ | ◆ | X | ○ | X |
| *Quandt, Grzywacz. 2014* | ● | ● | ● | X | X | X |
| *Solorio, Norton-Shelpuk. 2014* | ● | ○ | ● | ○ | ○ | X |
| *Song, Han. 2010* | ● | X | ● | X | ○ | X |
| *Suarez-Balcazar, Early. 2018* | X | ◆ | ◆ | X | ○ | X |
| *Vaughn, Jacquez. 2019* | ◆ | ◆ | ◆ | X | ○ | ◆ |
| *Wieland, Nigon. 2019* | ● | ● | ● | ● | X | X |
| *Wieland, Njeru. 2017* | ● | ○ | X | X | ○ | X |
| *Wieland, Wies. 2012* | ● | ○ | ● | X | X | X |
| *Williams, Ochsner. 2010* | ● | ◆ | ◆ | X | ○ | X |

◆ **Active participation:** Migrants (as the target population) were actively involved in decision-making processes and appeared to share power over decisions relating to this stage of the research.
● **Proxy-participation:** Evidence community groups/third-sector organisations and/or professional services were actively involved and wielded influence in decisions relating to this stage of the research. However, migrants (as the target population) did not appear to, and/or were indirectly participating.
○ **Indirect participation:** Evidence migrants (as the target population) were subject to research activities (e.g., surveys, interviews), or partook in implementing/actioning research directions (e.g., delivering the designed intervention), but did not appear to have any active involvement or influence in decisions relating to this stage of the research. Furthermore, there was no evidence of community group/third-sector organisation involvement or influence.
X **No explicit evidence:** There was no explicit evidence of the participation of migrants, community groups/third-sector organisations, or professional services regarding decision making processes related to this stage of the research stage.

However, there was greater variation and divergence in participatory research approaches when considering the degree of participation of migrants. In our analysis only 18 of the 28 included studies exhibit active participation of migrants (as the primary focus and target of the intervention) at any stage of the research process.^18,19,21-27,29-31,33,37,38,42-44^ Of these 18 studies, only two display evidence of active participation of migrants at all stages of the research process. ^25,26^

Across all 168 individual research stages analysed across the 28 studies, we recorded 46 instances of active participation of migrants; 30 instances of proxy participation; 24 instances of indirect participation; and 68 instances in which there was insufficient evidence to make a determination (Table 3). The active participation recorded also appears to be associated with the stage of the research. There were 7 instances of active participation during study inception; ^19,22,26,38,43-45^ 16 during design;^18,19,21-23,26,27,29-31,33,37,38,42,44,45^ 10 during implementation;^21,22,26,30,31,33,37,38,42,44,45^ 2 during analysis; ^26,45^ 6 during evaluation; ^24,26,30,31,44,45^ and 4 during dissemination. ^22,26,38,45^

### Benefits of participatory approaches to research

The benefit most often reported in utilising participatory approaches was the fact that interventions were better tailored to the target population through involving non-academic stakeholders. ^18,21,23,28,33,37,38,42,46^ This included two studies which spoke of the benefits of participatory research in facilitating interventions going beyond more immediately actionable cultural adaptations (such as language-adaptation and ethnically matched providers), to providing interventions that more deeply reflect community values and priorities. ^23,38^

Partnering with non-academic stakeholders was cited as providing practical benefits during the research. One study reported how participatory approaches allowed for the modification of the research programme throughout conception, development, and implementation. ^30^ Multiple publications provided evidence on how iterative feedback from stakeholder during the studies could aid partnerships, improving the recruitment of individuals to implement or take part in the intervention, ^17,18,23,28,38,47^ and dissemination. ^23^ One study also highlighted that partnerships were a feasible and appropriate means to support intervention implementation, ^20^ whilst one set of authors reported that partnerships with non-academics can ultimately strengthen research. ^22^

Better relationships between the community and academics were cited as having the capability to enhance the familiarity and trust of partners. One study cited that increased trust had direct benefits to research, leading to more open and honest dialogue than in traditional research, improving the accuracy and findings of these activities. ^43^ Researchers becoming part of ongoing community relations was seen as positive, or a catalyst, acting as an impartial bridge between disparate community groups. ^18^ Long-lasting partnerships built over the course of research was cited as producing a capacity building element, increasing the health-related knowledge and resources of the community which academics partnered with. ^24,31,39^ Finally, partnerships catalysed a greater degree of understanding of a subject among communities, leading to increased self-determination and the ability to generate change of their own accord. ^43^

### Challenges of participatory approaches to research with migrants

Multiple studies highlighted the importance of balancing the culture and expectations of both researchers and partnered individuals to enact participatory research. ^18,23,35^ For example, one study reported that reaching equitability in the research process and working on level-terms with migrants directly conflicted with the cultural norms of some partners, who revere authority figures, and so would in normal circumstances defer to their judgement. ^43^ A further study highlighted challenges exist in bringing together differing stakeholders, with varied views and experiences. In these situations it was suggested there is no ‘one size fits all’ approach, and that processes must be adapted to individual partner groups. ^31^ Noting varied perspective, one study highlights the challenge that the divergent perspectives of what is most salient and important to address amongst partners can present a challenge. ^25^

The challenge (and importance) of building rapport and addressing mistrust, ^18,19,44^ or even research fatigue amongst some partner groups, ^18^ was also evident within publications. One set of authors, spoke not just of the need for non-academic partners to trust researchers, but how It is also essential for researchers to trust wider stakeholder partners, and prioritise the collaborative and democratic aim of participatory methods; this was perceived as challenging as it may shift the power dynamic and locus of control in the research away from the academics. ^34^ Even when partnerships overcome challenges of culture, expectations and trust, there remain other practical challenges to operationalising these partnerships. ^29,33^

Challenges in ensuring equitability in research understanding, and balancing the participatory nature of a project, with the standards expected by the wider scientific community were also highlighted. ^18,31^ Furthermore, one study cited the difficulty of navigating acknowledgement and authorship of non-academics in published materials. ^23^ The lack of recognition of the requirements of participatory research in traditional academic circles is also cited as a challenge, with one set of authors stated the need for managerial, institutional, and funder-level buy-in and commitment regarding participatory research. ^18^ Similarly, institutional review limited the participation in at least one study, preventing non-academic stakeholders participating in data collection and analysis. ^31^

Other practical challenges to operationalising partnerships included effective, timely communication, ^19,32^ and the challenge of partnering with communities in which the dominant language of the researchers and migrant communities differ. ^35,43^ Finally, the iterative and tailored nature of the interventions produced may also impact on the generalisability of findings, ^37^ whilst some work could seemingly omit or contradict research evidence due to localising the intervention. ^40^

## Discussion

To our knowledge this is the first systematically review to robustly measure the application of participatory approaches and principles to health intervention research with migrants. Whilst specifically focusing on research with migrants, many of the findings, and the framework discussed are likely to provide insight to all practitioners of participatory research. The 28 studies included reported on a variety of health interventions, predominantly revolving around outreach and education. Our analysis shows that 18 of the 28 included studies actively involving migrants themselves, but only two studies actively involved migrants during all stages of the research process. The remaining studies either provide insufficient evidence to determine the participatory approach utilised, or were characterised by pseudo-participation, in which community groups/third-sector organisations were directly involved (proxy-participation), or migrants were only involved through being subjects in research activities (indirect-participation).

The approaches taken to participatory research in the studies included in this systematic review varied. The examples that represent the most participatory approach, according to our analysis, were characterised by co-operation and power-distribution (Table 4). The difference between active participatory approaches, and those we characterised as pseudo-participation, appear subtle when viewed from a research-centric perspective but are stark when considering a participatory perspective. Firstly, indirect participation, in which migrants are involved in activities such as surveys or interviews designed to inform health interventions may represent a perfectly suitable means to guide development and build evidence, but do little to distribute power in a participatory manner. The risk that research is framed as participatory whilst failing to develop equitable partnerships has previously been highlighted, and still appears to persist. ^3,48^ Secondly, proxy-participation, which may do more to uphold principles of participatory research, may still be at risk of not equitably involving the actual target population. Uncertainty persists around how to best involve non-academic stakeholders, and ensure those that are involved are representative of the population of interest. ^49^ Whilst community-groups and/or professional services involvement may at times be the only, or most readily available way to represent the population of interest (due to difficulties (perceived or otherwise) in accessing, or providing access to migrants), they cannot be assumed to be representative of them. Previous research has shown that health-service users can identify different needs to service-providers. ^50^ Furthermore, whilst overall understandings of involvement processes may align, service-providers may place different values on some aspects of involvement. ^51^ Therefore, proxy-participation could conceivably skew participatory research away from being centred on migrants’ needs.

**Table 4.** Descriptive tabulation of two studies classified as displaying active participation throughout all stage of the health intervention research with migrants.

|  |  | Research Stage |  |  |  |  |  |
| --- | --- | --- | --- | --- | --- | --- | --- |
|  |  | Inception | Design | Implementat<br>ion | Analysis | Evaluatio<br>n | Disseminati<br>on |
| Studi<br>es | Goodkin<br>d, Amer.<br>2017 | Study conceived from previous relations with community groups. The present study was guided by refugees, and community service providers. | The community was involved in designing interview protocols and participant recruitment procedures. | All interpreters and interviewers were refugees; procedures had been agreed during inception and design. | Refugees were involved in analysis and were actively involved in setting the agenda for what evidence was meaningful. | Refugees and community involved in discussions to evaluate the process; indication that the decision a community intervention paradigm be adopted appears to have been adopted and championed by researchers as a result. | Refugees were involved in the dissemination and are co-authors of the research publication. |
|  | Jacquez,<br>Vaughn.<br>2019 | Manifested from a previous relationship with latinos unidos por la salud to promote health and | co-researchers worked with academic partners to identify primary outcomes | Co-researchers recruited and worked with participants to identify strategies for stress reduction. | co-researchers and academic partners identified the primary outcomes . | Academic and community partners shared decision-making in all aspects of the | Academic and community partners shared decision-making in all aspects of the research |
|  |  | healthcare for the local latino community; co-researchers in this project were drawn from the local community. | , and helped decide that health-worker delivered strategies were the preferred intervention option |  |  | research process, including evaluation. | process, including dissemination. |

We found that involvement across all stages of the research was limited, and least prevalent in interpreting or disseminating research findings. Our findings are supported by and corroborate existing research, including two systematic reviews carried out to analyse how CBPR enhanced clinical trials for racial and ethnic minority groups, ^52^ and what it means to conduct participatory research with indigenous peoples. ^53^ These reviews found similar trends, including that participation varies substantially, and that research that is expressly participatory is often limited with key methodological challenges, around collecting, interpreting, and disseminating research. However, these reviews align with this research highlighting benefits of participatory research for the recruitment and retention of trial participants.

Upholding the core principles of participatory research, in this instance, democratising research and power-sharing, is particularly pertinent to partnering with migrants. Participatory research origins are firmly rooted in increasing social justice, and the promotion of doing research with, not on or about individuals and communities, particularly those that are disadvantaged. ^48^ Migrant communities are often marginalised within recipient countries, ^10,11^ and by local health systems. ^8,9^ Not only is it inappropriate for research to perpetuate or deepen any marginalisation through failing to include migrants’ voices, insights, and skills, but there are benefits to the proper utilisation of participatory approaches to the overall research process. Included studies provide evidence of the benefits to participant recruitment, implementation, and dissemination. Researchers also highlighted that the iterative nature of participatory research allows more effective tailoring of work to the needs of migrants, through learning from and embedding migrant partners knowledge and experience into research. Whilst studies we identify predominantly focus on community outreach and education within health research, participatory research could be better utilised across all disciplines. Similar methodology could be employed to better design pharmaceuticals, or on a larger scales, procedures and systems at a governance level.

Effectively partner with migrants requires specific strategies, to address the challenges identified in this review. Some of these strategies include: early participatory involvement to guide research priorities, methodological approaches, and strategies to manage ongoing relations; translating and back-translating materials; giving reassurance as to the confidentiality of involvement, and respecting decisions around reporting (particularly as some partners may be undocumented migrants or have precarious legal status); utilising a variety of outreach and recruitment outlets, such as NGOs and religious groups trusted by migrants; and identifying opportunities for bidirectional benefits in the research, and capacity building to facilitate collaborative and democratic participation. Those partnering with migrants must demonstrate flexibility to negotiate potential power divides, and acknowledge and be considerate of residual mistrust that may exist among communities, even after researcher-community relationships appear well established. ^54^ The challenges and extra consideration highlighted by this review must not be underestimated, whilst from a research perspective, more still needs to be done to assess the impact of participatory approaches on overall research processes and output. However, if research is to become more democratic, patient-centred, and representative of the populations impacted by its work, traditional scientific approaches are likely to be inadequate, with academic researchers holding the majority of power over research. ^55^

Greater adoption of consistent and transparent reporting of participatory research is needed, to support the need for more critical analysis of involvement and participatory research. ^56^ Whilst guidelines have been developed, ^57^ they have not been widely adopted, with no material improvement in the reporting quality of published studies seen within some fields as a result of the their publication, which could be attributed to a lack of awareness of the guidelines existence. ^58^ Tensions exist as to whether participatory research should be conceptualised and evaluated similarly to traditional research^56,59^ However, we believe reporting can be sympathetic of the need to evidence impact of methods and processes on research. We propose the plain reporting of who was involved in each element of the research, why these individuals were involved; how they were involved; and who ultimately controlled decisions relating to each research stages should be completed for all research involving non-academic stakeholders. The development of guidelines to support this reporting would simultaneously allow a more complete assessment of how participatory research impacted research outcomes, and greater reflection and evaluation of the participatory approaches employed. ^59^

Our review has several limitations. Firstly, we acknowledge the taxonomy of terminology around participatory research is not standardised and terms are used inconsistently. As such, the categorisations we have introduced and used in this review may be defined differently by others.. Furthermore, whether the included studies categorised in our analysis exhibit greater or lesser participation is potentially immaterial to the quality of the research carried out, or the impact of the final intervention. This review represents our attempt to cast a critical lens over how the principles of participatory research are applied in practice. Our conceptual focus on migrants’ involvement is therefore not intended to denigrate the efforts of third-sector organisations or professionals services, whose involvement we may have classified as proxy-participation. Fundamentally we believe the examined studies are inherently more participatory than traditional research endeavours, for having even considered and attempted to involve non-academic stakeholders. We recognise the challenges associated with participatory research, and as stated, hold no assumptions about the extent of participation and its’ association with beneficial outcomes for target populations.

## Conclusion

In conclusion, participatory approaches to research for health interventions aimed at migrants are insufficiently applied and reported. We provide evidence that the application of approaches does not fully embody core principles of participatory research, particularly relating to providing decision-making power to individuals ultimately affected by the research. Those who wish to engage in participatory research must consider the approach they take, and whether it is sufficient to achieve high-quality participation, not just high-quality research. Crucially, guidelines for reporting of participatory research methods must be introduced. This will enable all parties, from academics to communities to better assess the participatory nature of individual research projects, and is an important pre-requisite to exploring the overall impact of participatory research which is inadequately understood.

## Supporting information

Appendix 1. Representative search terms/strategy used

Summary of extracted data relating to involvement and engagement of non-academic stakeholders, and their influence over given research stages.

## Data Availability

All data generated or analysed during this study are included in this published article.

## Declarations

### Ethics approval and consent to participate

This research is a systematic review of existing literature. As such, no human participants were involved.

### Consent for publication

Not applicable

## Availability of data and materials

All data generated or analysed during this study are included in this published article.

## Competing interests

The authors declare that they have no competing interests.

## Funding Statement

This work has been supported by the European Society of Clinical Microbiology and Infectious Diseases (ESCMID) Study Group for Infections in Travellers and Migrants (ESGITM). KR is funded by the Rosetrees Trust (M775). SH, AFC, and LG are funded by the NIHR (NIHR Advanced Fellowship NIHR300072) and SH and AFC are funded by the Academy of Medical Sciences (SBF005\1111). AD is funded by the MRC (MR/N013638/1).

## Author Contributions

The study was conceptualised by KR, LN, SH and JSF with input on finalising the study question and protocol from SM-H and AC. Searches were carried out by KR, and screening done by KR, SM-H and AC. Data extraction and analysis was done by KR, AC and SH. Interpretation of the results and drafting of the manuscript was done primarily by KR with input from all authors. All authors commented upon and approved the final manuscript.

## Acknowledgements

Not applicable

## Authors’ information

Not applicable

## Additional Files

Additional file 1.docx – Representative search strategy and terms used in the systematic review. This additional file contains a table displaying a representative search strategy used to source articles through database searching.

Additional file 2.docx – Summary extracted data from studies included in this systematic review. This additional files contains a table displaying the summary extracted data obtained and analysed in this systematic review.

## Notes

### Competing Interest Statement

The authors have declared no competing interest.

### Author Declarations

As a systematic review of existing literature, this research did not involve human participants and did not require ethics approval

