## Appendix 1. Representative search terms/strategy used for "Participatory research in health intervention studies involving migrants: a systematic review"

**Migrant identifier terms**

Migrant* *or*

Migrat* *or*

Refugee* *or*

Foreign* *or*

Non-native* *or*

immigrant* *or*

emigra* *or*

oversea* *or*

Foreign students

**Participatory research terms**

Co-design* *or*

Co-prod* *or*

Co-creat* *or*

Collab* design* *or*

Collab* prod* *or*

Collab* create* *or*

Community design* *or*

Community prod* *or*

Community create* *or*

Community-based participatory research *or*

Action research *or*

Participatory action research *or*

Participatory adj4 research *or*

Patient involvement *or*

Patient and public involvement

Appendix 1. Representative search terms/strategy used
