## Supplementary material for "Participatory research in health intervention studies involving migrants: a systematic review": Summary of extracted data relating to involvement and engagement of non-academic stakeholders, and their influence over given research stages.

Appendix 2. Summary of extracted data relating to involvement and engagement of non-academic stakeholders, and their influence over given research stages.

1. **Afifi, Makhoul et al. 2011**

***Aim & Target population:*** *Development of a mental health promotion intervention for Palestinian refugee youths.*

***Inception:*** *Based on a previous survey of the target population which had been designed with community input*

***Design:*** *Interview and survey of target population; formation of a community youth council with youth council representatives which agreed internal rules and implemented democratic voting re: design decisions*

***Implementation:*** *No explicit evidence of non-academic stakeholder involvement, engagement, or influence.*

***Analysis:*** *No explicit evidence of non-academic stakeholder involvement, engagement, or influence.*

***Evaluation:*** *No explicit evidence of non-academic stakeholder involvement, engagement, or influence.*

***Dissemination:*** *No explicit evidence of non-academic stakeholder involvement, engagement, or influence.*

2. **Aitaoto, Braun et al. 2012**

***Aim & Target population:*** *Implementation of a cancer outreach programme for Micronesian women*

***Inception:*** *Community group representing the target population approached researchers to highlight this topic*

***Design:*** *Focus groups with the target population; champions from the target population were involved in literature review and aided in designing the outreach programme*

***Implementation:*** *Target population delivered the programme as lay-educators, a decision clearly defined in the design stage by working with the champions*

***Analysis:*** *No explicit evidence of non-academic stakeholder involvement, engagement, or influence.*

***Evaluation:*** *No explicit evidence of non-academic stakeholder involvement, engagement, or influence; reported that the research did not ask the target population about perceptions of the intervention*

***Dissemination:*** *No explicit evidence of non-academic stakeholder involvement, engagement, or influence.*

3. **Baird, Domian et al. 2015**

***Aim & Target population:*** *To address the health challenges associated with relocation among Sudanese refugees*

***Inception:*** *Collaborative inception between Sudanese community and researchers manifesting from previous research shared to community members, who then formed a committee which worked with the researchers; Whilst topics for intervention were collaboratively developed between researchers and the target population*

***Design:*** *research team developed educational seminars on intervention topics.*

***Implementation:*** *Educational seminars were delivered by the research team; target population were involved in advertising and recruiting other participants.*

***Analysis:*** *No explicit evidence of non-academic stakeholder involvement, engagement, or influence.*

***Evaluation:*** *Focus group discussion to evaluate the process, discuss overall findings, and plan future relationships.*

***Dissemination:*** *No explicit evidence of non-academic stakeholder involvement, engagement, or influence.*

4. **Barbee, Kobetz et al. 2010**

***Aim & Target population:*** *To assess the acceptability of HPV self-sampling with community health workers, to detect cervical cancer among Haitian immigrant women.*

***Inception:*** *Conceived by a campus-community partnership; the partnership is formed of community leaders and academics; community leaders are largely responsible for identifying research needs*

***Design:*** *Community partners responsible for defining research focus, recruitment strategies, and data collection methods; academic partners write grants applications and obtain institutional approvals*

***Implementation:*** *Community and academic partners contribute collectively to study implementation*

***Analysis:*** *No explicit evidence of non-academic stakeholder involvement, engagement, or influence.*

***Evaluation:*** *No explicit evidence of non-academic stakeholder involvement, engagement, or influence.*

***Dissemination:*** *Community and academic partners contribute collectively to study dissemination*

5. **Chesla, Chun. 2013**

***Aim & Target population:*** *Cultural adaptation of coping skills training for Chinese-American immigrants with type 2 diabetes.*

***Inception:*** *No explicit evidence of non-academic stakeholder involvement, or influence; there is mention of a CBPR work group, and a community advisory board, but no indication these groups were involved in the inception.*

***Design:*** *A CBPR work group composed of 10 paid staff from community agencies and academic institutions. The work group culturally adapted protocols based on clinical guidelines, clinical knowledge, and previous studies.; A community advisory board met semi-annually and provided oversight on all aspect of the CBPR activities.*

***Implementation:*** *Work group members who were bilingual with degree level qualification in social work implemented the intervention.*

***Analysis:*** *No explicit evidence of non-academic stakeholder involvement, engagement, or influence.*

***Evaluation:*** *Evaluated through research/survey with participants.*

***Dissemination:*** *No explicit evidence of non-academic stakeholder involvement, engagement, or influence.*

6. **Evans, Suggs et al. 2019**

***Aim & Target population:*** *Promoting HIV testing uptake amongst African migrant communities*

***Inception:*** *No explicit evidence of non-academic stakeholder involvement, engagement, or influence.*

***Design:*** *The design was guided by previous formative research conducted by the researchers*

***Implementation:*** *Participants were invited by community researchers that represented diverse African communities and genders.*

***Analysis:*** *No explicit evidence of non-academic stakeholder involvement, engagement, or influence.*

***Evaluation:*** *The target population provided feedback through surveys*

***Dissemination:*** *No explicit evidence of non-academic stakeholder involvement, engagement, or influence.*

7. **Forst, Ahonen et al. 2013**

***Aim & Target population:*** Increase awareness of workplace hazards and self-efficacy among foreign-born Hispanic construction workers; and expand worker centre agenda to include occupational health & safety.

***Inception:*** *No explicit evidence of non-academic stakeholder involvement, engagement, or influence.*

***Design:*** *Worker centres (WC) and academics partnered to set joint goals and specific responsibilities: WC would recruit peer-educators and workers to the training, partner with implementation and evaluation; university partners assumed responsibility for subject protection, evaluation, design and implementation.*

***Implementation:*** *Worker centre staff were responsible for recruiting peer-educators to deliver the curriculum; university partners assumed responsibility for implementation*

***Analysis:*** *No explicit evidence of non-academic stakeholder involvement, engagement, or influence.*

***Evaluation:*** *Iterative evaluation of the intervention with peer-educators in a democratic manner; peer-educators were able to influence study design*

***Dissemination:*** *No explicit evidence of non-academic stakeholder involvement, engagement, or influence.*

8. **Goodkind, Amer et al. 2017**

***Aim & Target population:* to address social determinants of health, improve links to mental health services, and improve retention in trauma-focused treatment for Afghan, Great lakes region african, and iraqi Refugee adults**

***Inception: Inception came from previous work with other community groups; present study guided by refugees, formers students and community service providers***

***Design: Community involved in designing the interview protocols, and participant recruitment***

***Implementation: target population involved in implementation; all interpreters and interviewers were refugees***

***Analysis: target population involved in analysis; active involvement in asking what evidence is meaningful***

***Evaluation: CAC were involved in conversation concerning the use of CAC; have voice opinions that a community intervention paradigm should be used which the researchers appear to have heeded.***

***Dissemination****: target population involved in dissemination; 2nd and 8th author are refugees*

9. **Grigg-Saito, Och et al.**

***Aim & Target population:*** *Outreach to reduce health disparities in cardiovascular disease and diabetes among Cambodian refugees*

***Inception:*** *Coalition formed prior to this intervention to address needs of Cambodian community - resulted in the funding bid for this work; the coalition involved community leaders and elders*

***Design:*** *Collaboration amongst the coalition to develop a community action plan. Target population took part in focus groups; community forum involving target population (engagement event) took place*

***Implementation:*** *Cambodian community health programme Staff health educators go door-to-door, staff also encouraged participation from elders; Staff conducted business outreach and educational groups/workshops; A peer-support group was established; elders council (advisory group) established at the behest of elder program participants*

***Analysis:*** *No explicit evidence of non-academic stakeholder involvement, engagement, or influence.*

***Evaluation:*** *Data was reported back to staff and steering committee periodically, everyone was able to contribute to lessons learned, draw conclusions and make suggestions for improvements based on the evaluation.*

***Dissemination****: No explicit evidence of non-academic stakeholder involvement, engagement, or influence.*

10. **Henderson, Slater**

***Aim & Target population:* Nutrition information for newly arrived immigrants**

***Inception: based on previous community-based participatory research***

***Design:*** ***Community consultation***

***Implementation:*** *No explicit evidence of non-academic stakeholder involvement, engagement, or influence.*

***Analysis:*** *No explicit evidence of non-academic stakeholder involvement, engagement, or influence.*

***Evaluation: community consultation on evaluation***

***Dissemination****: No explicit evidence of non-academic stakeholder involvement, engagement, or influence.*

11. **Jacquez, Vaughn et al. 2019**

***Aim & Target population:*** *Implementation of a stress reduction intervention for latino immigrants.*

***Inception:*** *Manifested from a previous relationship with latinos unidos por la salud to promote health and healthcare for the local latino community; community members applied and were selected as co-researchers*

***Design****: Community group co-researchers worked with academic partners to identify primary outcomes, and that health-worker delivered strategies were the preferred intervention option.*

***Implementation:*** *Co-researchers recruited and worked with participants to identify strategies with participants.*

***Analysis:*** *co-researchers and academic partners identified the primary outcomes*

***Evaluation:*** *Academic and community partners shared decision-making in all aspects of the research process, including evaluation.*

***Dissemination****: Academic and community partners shared decision-making in all aspects of the research process, including dissemination.*

12. **Kaiser, Martinez et al. 2015**

***Aim & Target population:*** *Adaptation of a culturally relevant nutrition and exercise program for Mexican immigrant parents with young children*

***Inception:*** *Researchers conceived through reviewing literature and involving the target population in focus groups to assess their interpretation and prioritisation of key childhood obesity messages.*

***Design:*** *Researchers performed a literature search and involved the target population in focus groups; the researchers presented the plans for a nutrition program to a local advisory committee whom convened quarterly and gave advice on how to deal with specific issues*

***Implementation:*** *A local educator with experience of family counselling was hired to deliver the intervention*

***Analysis:*** *No explicit evidence of non-academic stakeholder involvement, engagement, or influence.*

***Evaluation:*** *pilot testing with a subsection of families who were invited to give feedback; educator and specialist would evaluate intervention events after each one for changes; participants were invited to focus groups to report back on the intervention.*

***Dissemination****: No explicit evidence of non-academic stakeholder involvement, engagement, or influence.*

13. **Kandula, Dave et al. 2016**

***Aim & Target population:*** *Exercise intervention for south-asian mother with risk factors for diabetes*

***Inception:*** *Local community group and researchers collaboratively chose the focus of the research*

***Design:*** *Community partner and academics adapted and applied evidence-based behaviour change principles in partnership; a memorandum of understanding was agreed by the partners: The community group was involved in design, implementation, recruitment and evaluation.*

***Implementation:*** *Community partner was responsible for recruitment and outreach*

***Analysis:*** *No explicit evidence of non-academic stakeholder involvement, engagement, or influence.*

***Evaluation:*** *The community partner was involved in evaluation; participants were involved in the evaluation of the intervention through qualitative interviews.*

***Dissemination****: No explicit evidence of non-academic stakeholder involvement, engagement, or influence.*

14. **Karasz, Raghavan et al. 2015**

***Aim & Target population:*** *Development of an asset-building mental health intervention for Bangladeshi immigrant women*

***Inception:*** *Outgrowth from previous partnerships; the development involved local physicians, a health advocate and researchers.*

***Design:*** *The target population were included as part of the intervention design group*

***Implementation:*** *No explicit evidence of non-academic stakeholder involvement, engagement, or influence.*

***Analysis:*** *No explicit evidence of non-academic stakeholder involvement, engagement, or influence.*

***Evaluation:*** *No explicit evidence of non-academic stakeholder involvement, engagement, or influence; target population were involved in the evaluation through research methods.*

***Dissemination:*** *No explicit evidence of non-academic stakeholder involvement, engagement, or influence.*

15. **Kim, Koniak-Griffin et al. 2004**

***Aim & Target population:*** *Cardovascular health promotion among latino immigrants*

***Inception:*** *Collaboration with local department of health and nurses*

***Design:*** *Local department of health refined area for targeting; 2 community based organizations and the formation of an advisory board (including migrants) was instrumental in tailoring the study to the target community (e.g. changing name from lay health advisor to promotoras de salud).*

***Implementation:*** *Lay health advisors were recruited and trained, by the local department of health, to deliver the intervention*

***Analysis:*** *No explicit evidence of non-academic stakeholder involvement, engagement, or influence.*

***Evaluation:*** *Feedback from the local health department, community advisory boards and lay health educators allowed flexible iteration of the intervention*

***Dissemination:*** *No explicit evidence of non-academic stakeholder involvement, engagement, or influence.*

16. **Lam, McPhee et al. 2003**

***Aim & Target population:*** *Increase pap/cervical cancer screening among Vietnamese-american women*

***Inception:*** *No explicit evidence of non-academic stakeholder involvement, engagement, or influence.*

***Design:*** *Shared with the REACH (community) coalition (who had overall power); community forum was held with 200 vietnamese-american women and their families to discuss cervical cancer and brainstorm strategies to increase screening (the REACH coalition then took findings to generate the intervention); The results were shared with a second community forum, with feedback used to finalise the intervention (by the coalition).*

***Implementation:*** *The lay health worker campaign was community-led;* *Lay health educators were the target population; the community coalition was involved in implementing a media campaign.*

***Analysis:*** *No explicit evidence of non-academic stakeholder involvement, engagement, or influence; institutional review board deemed partners could not collect data.*

***Evaluation:*** *Media campaign was evaluated at a community forum in which community members opinions were sought through questionnaires and discussions. The REACH coalition retained the power to effect changes; lay health worker opinion was solicited through research methods/questionnaires; No evidence of individual health workers being able to influence intervention in progress.*

***Dissemination:*** *No explicit evidence of non-academic stakeholder involvement, engagement, or influence.*

17. **Li, Yeh et al. 2019**

***Aim & Target population:*** *Colorectal cancer prevention among Chinese-americans*

***Inception:*** *Researchers and leaders from community groups agreed to promote colorectal cancer prevention in Chinese-americans.*

***Design:*** *Designed between the researchers with feedback from the community group; Researcher carried out a literature review with feedback from community partner, research with the actual community in the form of qualitative interviews was conducted to inform the design; Appears community group did have influence, changed from a web-based idea due to community group input.*

***Implementation:*** *Delivered by the researchers*

***Analysis:*** *No explicit evidence of non-academic stakeholder involvement, engagement, or influence.*

***Evaluation:*** *The community group members were involved in filling out surveys as part of the evaluation*

***Dissemination:*** *No explicit evidence of non-academic stakeholder involvement, engagement, or influence.*

18. **Nilvarangkul, McCann et al. 2011**

***Aim & Target population:*** *Enhancing health-related quality of life for Laotian migrants*

***Inception:*** *Conceived by researchers with a broad action plan developed in collaboration with the target population*

***Design:*** *The target population were involved in interviews used to shape the intervention; meetings were held with community workers and stakeholders to create action plans*

***Implementation:*** *Community workers, stakeholders and researchers implemented the action plan*

***Analysis:*** *No explicit evidence of non-academic stakeholder involvement, engagement, or influence.*

***Evaluation:*** *Meetings were held with community workers and stakeholders as part of the formative evaluation; intervention participants were followed up via qualitative data collection*

***Dissemination:*** *No explicit evidence of non-academic stakeholder involvement, engagement, or influence.*

19. **Pinsker, Call et al. 2017**

***Aim & Target population:*** *Tobacco use prevention videos for Somali youth*

***Inception:*** *No explicit evidence of non-academic stakeholder involvement, engagement, or influence.*

***Design:*** *There was indirect involvement through focus groups of the target population; there was direct involvement of the target population in developing the videos, building upon the focus groups.*

***Implementation:*** *Target population were involved in shaping the implementation of the intervention*

***Analysis:*** *No explicit evidence of non-academic stakeholder involvement, engagement, or influence.*

***Evaluation:*** *Target population evaluated the intervention through participant interviews*

***Dissemination:*** *No explicit evidence of non-academic stakeholder involvement, engagement, or influence.*

20. **Quandt, Grzywacz et al. 2013**

***Aim & Target population:*** *Pesticide safety intervention for migrant farmworkers*

***Inception:*** *Based on an existing relationship between the university and community partners*

***Design:*** *Based on an existing partnership; not immediately obvious how taget population and non-academic stakeholders were involved.*

***Implementation:*** *Implemented by the community partners; lay health promoters were recruited from within the community and trained by the community partner*

***Analysis:*** *No explicit evidence of non-academic stakeholder involvement, engagement, or influence.*

***Evaluation:*** *No explicit evidence of non-academic stakeholder involvement, engagement, or influence.*

***Dissemination:*** *No explicit evidence of non-academic stakeholder involvement, engagement, or influence.*

21. **Solorio, Norton-Shelpuk et al. 2014**

***Aim & Target population:*** *HIV prevention messages for young Latino immigrant MSM*

***Inception:*** *There is a previous community-university relationship, uncertain as to where the inception for the project came.*

***Design:*** *Storyboard/intervention was put before the target group in focus groups, Latino MSM were hired to facilitate the focus groups and help interpret the findings*

***Implementation:*** *Intervention was actioned by an external digital marketing/media team who used the design focus groups to develop the campaign*

***Analysis:*** *Latino MSM were hired to help interpret the findings*

***Evaluation:*** *The target population were involved in focus groups to assess their comprehension of the intervention*

***Dissemination:*** *No explicit evidence of non-academic stakeholder involvement, engagement, or influence.*

22. **Song, Han et al. 2010**

***Aim & Target population:*** *Translating dietary guidelines into a culturally tailored nutrition education program for Korean-American immigrants with type 2 diabetes*

***Inception:*** *There is a previous relationship between researchers and community, but no clear link as to how this explicitly influenced inception*

***Design:*** *No explicit evidence of non-academic stakeholder involvement, engagement, or influence.*

***Implementation:*** *No explicit evidence of non-academic stakeholder involvement, engagement, or influence; appears the education program was delivered by nurses and dieticians*

***Analysis:*** *No explicit evidence of non-academic stakeholder involvement, engagement, or influence.*

***Evaluation:*** *Target population were involved in evaluating the interventions’ acceptability*

***Dissemination:*** *No explicit evidence of non-academic stakeholder involvement, engagement, or influence.*

23. **Suarez-Balcazar, Early et al. 2018**

***Aim & Target population:*** *Healthy lifestyle promotion among Latino immigrant families with youth with disabilities*

***Inception:*** *No explicit evidence of non-academic stakeholder involvement, engagement, or influence.*

***Design:*** *Focus groups with the target population; interviews and conversations with community stakeholders and agency personnel; research built on by collaboration with mothers and staff from community organisations*

***Implementation:*** *Delivered by bilingual occupational therapy and nutrition students; co-facilitated by mothers who presented Zumba sessions and prepared healthy snacks.*

***Analysis:*** *No explicit evidence of non-academic stakeholder involvement, engagement, or influence.*

***Evaluation:*** *Evaluated through focus groups at the end of the intervention*

***Dissemination:*** *No explicit evidence of non-academic stakeholder involvement, engagement, or influence.*

24. **Vaughn & Jacquez. 2019**

***Aim & Target population:*** *Stress and coping intervention for Latino immigrants*

***Inception:*** *There are existing relations between the researchers and community; migrant community members were selected to be co-researcher*

***Design:*** *Design input came from researchers and community co-researchers*

***Implementation:*** *Community co-researchers implemented the intervention*

***Analysis:***

***Evaluation:*** *Co-researchers provided evaluation through interviews with the researchers*

***Dissemination:*** *No clear evidence, though does mention co-researchers want to be involved in dissemination including publications and presentations*

25. **Wieland, Nigon. 2019**

***Aim & Target population:*** *Tuberculosis screening for migrant adults*

***Inception:*** *Conceived and initiated by a community organisation*

***Design:*** *Academic and non-academic community organisation co-designed the intervention; not sure of the involvement of the target population*

***Implementation:*** *Intervention was implemented by community organisation staff, along with clinical/nursing staff and students*

***Analysis:*** *Evidence was collected and analysed by local health practitioners*

***Evaluation:*** *No explicit evidence of non-academic stakeholder involvement, engagement, or influence.*

***Dissemination:*** *No explicit evidence of non-academic stakeholder involvement, engagement, or influence.*

26. **Wieland, Njeru et al. 2017**

***Aim & Target population:*** *Digital storytelling intervention for immigrants and refugees with diabetes*

***Inception:*** *Impetus came from an existing researcher-community partnership; diabetes co-agreed as an area for research and intervention*

***Design:*** *survey developed by community-academic partnership to understand target population, attitudes and behaviours in T2DM; focus groups were conducted to further inform intervention content and to identify gifted storytellers; storytellers were recruited by community partners and their stories were captured, recorded and edited to derive the final intervention product.*

***Implementation:*** *No explicit evidence of non-academic stakeholder involvement, engagement, or influence.*

***Analysis:*** *No explicit evidence of non-academic stakeholder involvement, engagement, or influence.*

***Evaluation:*** *participants completed follow-up surveys*

***Dissemination:*** *No explicit evidence of non-academic stakeholder involvement, engagement, or influence.*

27. **Wieland, Wies. 2012**

***Aim & Target population:* Physical activity and nutrion program for immigrant and refugee women**

***Inception: Conceived from a community-academic partnership with long-standing ties; formalised to use CBPR to improve migrant health***

***Design: Exercise programme piloted; refugee and immigrant women were included as focus group participants***

***Implementation: Implemented by researchers and community organisation; community organisation recruited women to the programme; community organisation provided facilities***

***Analysis:*** *No explicit evidence of non-academic stakeholder involvement, engagement, or influence.*

***Evaluation:*** *No explicit evidence of non-academic stakeholder involvement, engagement, or influence.*

***Dissemination:*** *No explicit evidence of non-academic stakeholder involvement, engagement, or influence.*

28. **Williams, Ochsner et al. 2010**

***Aim & Target population:*** *Health and safety training for Latino day laborers*

***Inception:*** *Inception appears to come from advocacy/community groups*

***Design:*** *Focus groups held with the target population to identify typical exposures and situations to be addressed by peer trainers; curriculum was revised with feedback from these peer trainers*

***Implementation:*** *Delivered by peer-trainers*

***Analysis:*** *No explicit evidence of non-academic stakeholder involvement, engagement, or influence.*

***Evaluation:*** *Evaluated through focus group discussions and pre & post-training surveys;*

***Dissemination:*** *No explicit evidence of non-academic stakeholder involvement, engagement, or influence.*
